# Factors associated with delayed access of care among children under five with malaria and their outcomes at a regional referral hospital in Eastern Uganda

**DOI:** 10.64898/2026.07.16.26358296

**Authors:** Kayeny Miriam Melody Yung, Lydia VN Ssenyonga, Paul Oboth, Lyagoba Ivan, Samuel Olowo, Pamella R. Adongo

**Affiliations:** Department of Nursing, Busitema University Faculty of Health Sciences, Uganda, P.O Box 1460; Department of Community and Public Health, Faculty of Heath Sciences, Busitema University, P.O. Box 1460, Mbale, Uganda

## Abstract

**Introduction:** Uganda has the highest number of Malaria cases in East and Southern Africa and accounted for 3% of deaths in 2020. This calls for prevention, early diagnosis and treatment of Malaria most especially among children whose condition can progress to severe Malaria within 24 hours. While numerous interventions have been put into place to prevent malaria transmission, delays in diagnosis and treatment of Malaria when ill can lead to further mortality. Therefore, factors associated with delayed access of care among children under five with malaria and their outcomes need to be explored.

**Methods:** A cross sectional study was carried out. The target population was parents/caretakers to children under five with malaria at Mbale Regional Referral Hospital. A consecutive sampling technique was used on the target population. Quantitative data was collected using researcher administered questionnaires designed consistent with the research objectives. Collected data was analyzed using STATA version 15.

**Results:** Among the 216 children under five admitted at Mbale regional referral hospital with Malaria, 59.26% received care from a health facility 24 hours after symptom onset. The most significant predictors of delay in seeking care were the caregiver/ parent having attained tertiary education (AOR=7.1, p value=0.02) and initially implementing other measures other than giving medication/herbs before taking a child to the health center (AOR=4.1, p value=0.00).

**Conclusion:** Despite the numerous interventions put into place to curb the spread of malaria and to manage malaria, delayed access of care remains a significant contributor to the adverse effects of malaria among children under five. Health education on the impact of delayed access of care should be intensified at all levels of healthcare.

## Introduction

Delay is a period of time when someone/something has to wait because of a problem that makes something slow(1). In regards to health and disease progression, delays increase the risk of developing complications(2).

Malaria in young children can progress to a severe form within 24 hours after the onset of symptoms(3). Malaria is an acute febrile illness caused by plasmodium parasites which are spread to people through bites of infected female anopheles mosquitoes(4). In East and Southern Africa, Uganda has the highest number of malaria cases with a prevalence of 23.7 %(5). Between 2018 and 2019, 15% of the children under five tested for malaria in Bugisu region had malaria (6). The above data indicates that malaria is major burden of disease among children especially those under five and needs prompt interventions.

Appropriate and efficacious medication needs to be administered on time(7). This however isn’t the case due to delays that occur in access of care(7). Different studies show that these delays are associated with traditional beliefs(8), severity of signs(9), the structure of the healthcare system, the use of herbs before seeking professional care(10), various responsibilities of caregivers that make them give less priority to ill health(11), proximity to the health facility(12), low economic status(13), the perception that malaria was just a common disease(14) and the parent/caregiver’s highest level of education(15). Various factors are associated with these delays however no published study has been carried out to ascertain their causes and outcomes at Mbale Regional Referral Hospital, hence the relevance of this study.

## Methods and Materials

### Study Design

A cross sectional study design was used to obtain information about the factors associated with delayed access of care among children under five with malaria and their outcomes at one specific time. Cross sectional studies are descriptive and allow one to observe many variables at one point in time using many study participants. Quantitative methods were employed using researcher administered questionnaires that were administered to parents/caretakers of children U5 with malaria at MRRH.

### Study Area

This study was carried out at Mbale Regional Referral Hospital (MRRH) in Mbale District, Eastern Uganda. MRRH is located along Pallisa road in Mbale city. MRRH is the referral hospital for the districts of Busia, Budaka, Kibuku, Kapchorwa, Bukwo, Butaleja, Manafwa, Mbale, Pallisa, Sironko and Tororo among others. MRRH also however serves many more patients outside the hospital’s catchment area. MRRH is a public hospital with a capacity of about 470 beds and serves a population of nearly 4.5 million people. MRRH provides promotive, curative, palliative and rehabilitative health services. From J ^MRRH^ to May 2022, there were 7,424 admissions due to Malaria among children under five at MRRH. MRRH’s pediatrics unit has three pediatricians, one pediatric nurse (SPNO MRRH) and a 36 bed capacity.

**Fig 1.**
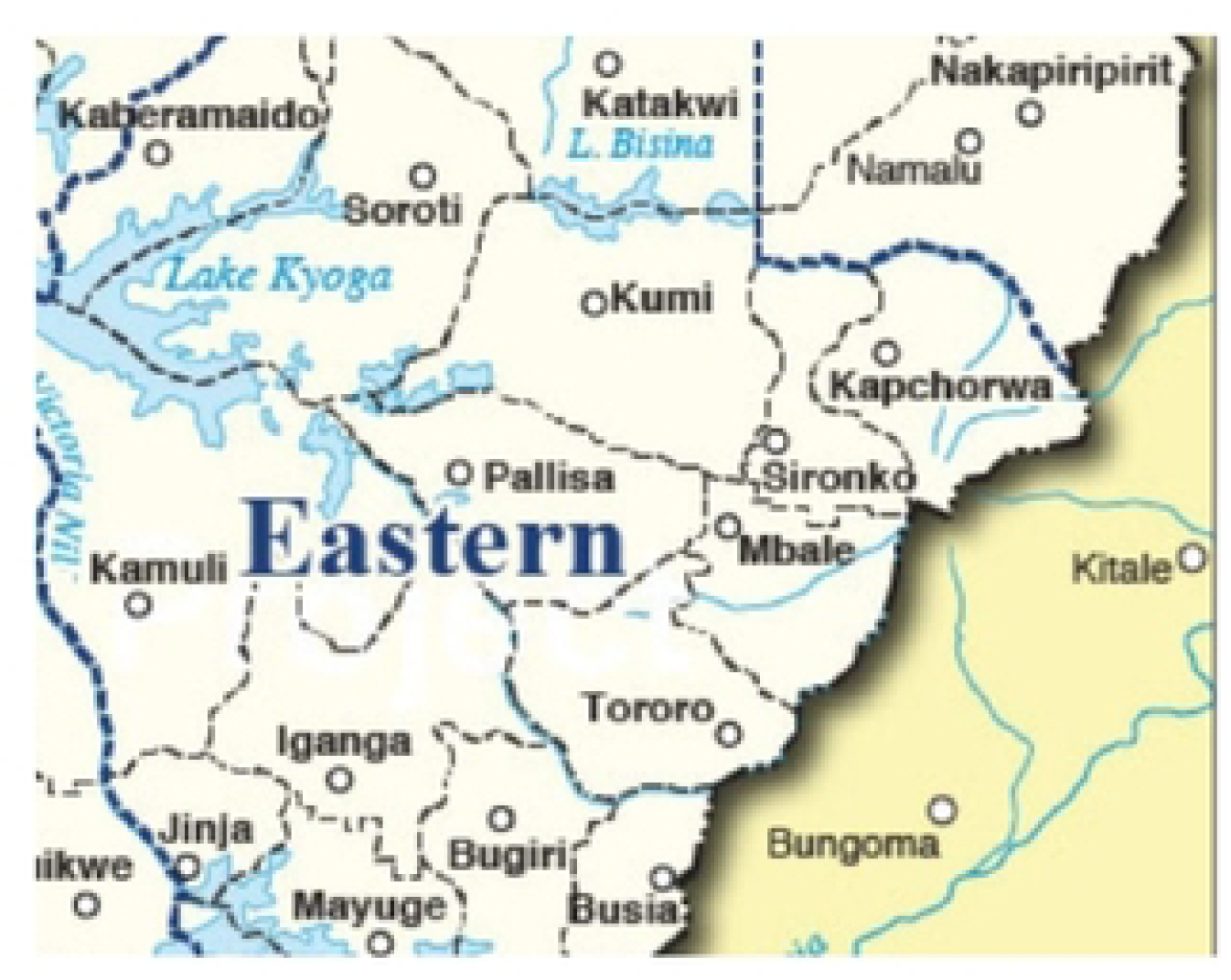
A map of Eastern Uganda showing Mbale Regional Referral Hospital (MRRH) and the neighboring districts

### Study Population

The study included parents/ caretakers to children under five with malaria admitted at MRRH when data was being collected.

### Sample Size

The sample size was calculated using Fisher’s formula,

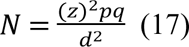

Where;

### N was the sample size

z was the standard normal deviation (1.96) corresponding to 95% confidence interval p was 15% which is the prevalence of malaria in Bugisu among children U5(6)

d was the degree of accuracy (+/-5%)

q= (1-p)

Therefore, N = [(1.96)2 x 0.15x 0.85] / (0.05)2

= 196 participants.

With a 10% non-response rate, the sample size was (196+20)

= 216 participants.

### Sampling Procedure

A consecutive sampling plan was used. Here, the researcher included all those who met the inclusion criteria and were conveniently available. This sampling procedure is useful for exploratory studies since it avails sufficient information for the study.

### Eligibility Criteria

#### Inclusion criteria

Parents/Caretakers to children U5 having malaria who consented to participate in this study at MRRH were included in this study.

#### Exclusion criteria

Mentally incapacitated, critically ill, deaf and dumb parents/caretakers to children U5 having malaria were excluded from this study since they were perceived to be unable to provide the relevant information.

### Study Variables

#### Independent variables

Factors associated with delayed access of care among children U5 with malaria. The factors were categorized under decision making, getting to the health facility and receiving adequate care at the health facility. Factors under decision making included the age, sex, education level, occupation and marital status of the parent/ caregiver. Factors under getting to the facility included the distance to the health facility, transport means and transport costs. Factors under receiving adequate care at the health facility included waiting time, availability of drugs and attitude of the health worker.

#### Dependent variable

The delay in access of care. Delay is taking more than one day/ twenty four hours to seek professional care after the onset of signs/symptoms of malaria. In this study, parents/caretakers to children U5 with malaria were considered to have delayed if they took more than 24hours to seek professional medical care. The delay then leads to outcomes like severe malaria, prolonged hospital stay and recovery. Prolonged hospital stay is a hospital admission of more than 5 days with a confirmed severe malaria diagnosis.

#### Data Collection

An electronic data collection and storage tool in form of Google forms accessible on phones and laptops from which researcher administered questionnaires were used to collect data. The interviews were carried out from the acute and pediatric wards at MRRH and each lasted 20 minutes.

#### Pre-testing the questionnaire

If the prevalence of the problem was 0.20, 8 participants were necessary to achieve a power of 80 % (18). The questionnaire was pre- tested on ten parents/caretakers to children U5 with malaria at Soroti Hospital to assess its reliability and to know the average amount of time required to collect the data. These ten parents/caretakers were not considered to be part of the study.

#### Data sources

An electronic data collection and storage tool in form of Google forms accessible on phones and laptops from which researcher administered questionnaires were used to collect data. The interviews were carried out from the acute and pediatric wards at MRRH and each lasted 20 minutes. Information provided by caretakers was verified against patients’ medical records to ensure accuracy and consistency.

### Data Management

#### Data collection and storage

The raw data was collected using questionnaires and stored in a computer data base tool and access was limited to the principal researcher. Data collection commenced on 27/10/2022 and ended on 30/11/2022. To address the challenge of language barrier, two nurses who were well conversant with local languages like lugisu, ateso and lugwere were selected to help in translation of the data collection tool to the parents or caretakers.

#### Data Analysis

Data collected was analyzed using STATA version 15. Categorical variables were summarized as proportions and continuous variables as means and standard deviation. Bivariate regression was used to show how an independent variable predicted the dependent variable. Factors with a p value less than 0.05 at bivariable analysis were considered to be significant and were then included in the multivariable analysis. Multivariate logistic regression was used to determine factors associated with delay in access of care among children with malaria and their outcomes while controlling confounders. A backward elimination method was used to get statistically significant variables. Factors with a p value less than 0.05 at multivariable analysis were considered to be significant.

#### Data Presentation

The analyzed data was presented in form of statistical tables, charts and graphs with interpretive descriptions of the information. This was done to ease the understanding of the data collected and to show relationships between variables.

#### Data Dissemination

The findings of this study were disseminated to Busitema University faculty of health sciences and MRRH.

#### Ethical Approval and Consent

Ethical procedures were all met, with approval obtained from Mbale Regional Referral Research and Ethics Committee (UG-REC-011) under the reference number MRRH-2022-208. Upon approval, participants were recruited and data collected. Because the study involved human subjects, informed consent was obtained from all participants prior to their involvement. Written consent forms were provided to those who were able to read and write. For participants who could not read or write, the consent form was read aloud in a language they understood, and consent was documented through a thumbprint in the presence of a witness. Parents/caretakers below the age of 18 were considered as emancipated minors and eligible to take part in the study. Ethical principles were adhered to throughout the research process in accordance with the law and institution guidelines.

## Results

### Socio-demographics characteristics

A total of 216 respondents took part in this study.

Socio-demographic characteristics showed that more females (74%) took part in the study and that 64.35% of the respondents were between the ages of 18 and 35 years with a mean age of 33 years (SD ±10).

For the rest of the other socio-demographic characteristics of the respondents, refer to the table below.

**Table 1:** Socio-demographic characteristics of the respondents.

| VARIABLE |  | FREQUENCY | PERCENTAGE (%) |
| --- | --- | --- | --- |
| Age | <=18 years | 5 | 2.31 |
|  | >18 to 35 years | 139 | 64.35 |
|  | >35 to 50 years | 60 | 27.78 |
|  | Above 50 years | 12 | 5.56 |
| Gender | Female | 160 | 74 |
|  | Male | 56 | 26 |
| Marital status | Married | 180 | 83.33 |
|  | Single | 16 | 7.40 |
|  | Widowed | 18 | 8.33 |
|  | Divorced | 12 | 5.60 |
| Highest level of education | No formal education | 24 | 11.11 |
|  | Not completed primary education | 82 | 38 |
|  | Completed primary education | 50 | 23.11 |
|  | Not completed secondary education | 42 | 19.44 |
|  | Completed secondary education | 5 | 2.31 |
|  | Tertiary education | 13 | 6.01 |
| Occupation | Farmer/casual laborer | 149 | 68.98 |
|  | Salaried | 18 | 8.33 |
|  | Student | 6 | 2.78 |
|  | Business man/woman | 43 | 19.91 |
| Sex of the child | Male | 123 | 56.94 |
|  | Female | 93 | 43.06 |
| Housing | Permanent house | 58 | 26.85 |
|  | Semi-permanent | 118 | 54.63 |
|  | Temporary | 40 | 18.52 |
| Number of children in the household | 1 to 4 | 122 | 56.48 |

### Summary of the delays

Majority of the respondents (69.44%) had children who had been referred from peripheral health facilities to MRRRH for further management. Most of the respondents (59.26%) took more than 24 hours to reach the health facility when their children fell ill. The average time taken to seek care was 1 day and 6 hours.

The other factors associated with the delay to reach the health facility and delays in the health facility are summarized in the table below.

**Table 2:** Factors associated with the delay to reach the health facility and delays in the health facility.

| VARIABLE |  | FREQUENCY | PERCENTAGE (%) |
| --- | --- | --- | --- |
| Referral system | Self-referral | 66 | 30.56 |
|  | From peripheral facility | 150 | 69.44 |
| When the child was not feeling well, what did you do | None | 12 | 5.56 |
|  | Herbs | 8 | 3.70 |
|  | Gave medication | 133 | 61.57 |
|  | Others | 62 | 28.70 |
| Where did you first go when the child started feeling unwell | Drug shop/ pharmacy | 64 | 29.63 |
|  | Private clinic | 50 | 23.15 |
|  | Public health facility | 63 | 29.17 |
|  | MRRH | 35 | 16.20 |
|  | Others | 4 | 1.85 |
| Transport means | On foot | 25 | 11.57 |
|  | Motorcycle | 170 | 78.70 |
|  | Bicycle | 8 | 3.70 |
|  | Car | 13 | 6.02 |
| How far is your home from the nearest health facility | <=500m | 73 | 33.80 |
|  | >500m to 2km | 97 | 44.91 |
|  | >2km | 46 | 21.30 |
| Time taken to reach the initial health facility when the child was ill | <24hrs | 88 | 40.74 |
|  | >=24hrs to 3 days | 111 | 51.39 |
|  | >3 days | 17 | 7.87 |
| If you spent more than 24 hours, what other factors contributed to this? | Did not have enough money | 53 | 24.54 |
|  | Waited for the partner to decide | 8(3.70) | 3.70 |
|  | Previous negative encounter with healthcare system | 1 | 0.46 |
|  | Thought the child would improve | 94 | 43.52 |
|  | Others | 22 | 10.19 |
| How long did it take to meet the health worker at the initial health facility? | <= 30 minutes | 162 | 75 |
|  | >30 minutes to 3 hours | 32 | 14.81 |
|  | >3 hours to 1 day | 22 | 10.19 |
| How long did it take to receive the initial treatment? | Less than 30 minutes | 69 | 31.94 |
|  | More than 30 minutes but less than 24 hours | 144 | 66.67 |
|  | More than 24 hours | 1 | 0.46 |
|  | None of the above | 2 | 0.93 |
| Were the prescribed drugs available at the initial health facility? | Yes | 84 | 38.89 |
|  | No | 132 | 61.11 |
| If your answer was “no”, would this stop you from returning to the health facility for health services? | Yes | 16 | 10 |
|  | No | 119 | 90 |
| Health worker’s attitude | Friendly and resourceful | 194 | 89.81 |
|  | Rude and unapproachable | 20 | 9.26 |
|  | Others | 2 | 0.93 |
| If your answer was “rude and unapproachable”, would that stop you | Yes | 6 | 30 |
| from returning to the health facility for health services |  |  |  |
|  | No | 14 | 70 |

### The initial management of a child with malaria while at home

Most of the parents/ caretakers (61.57%) reported to having given medication to their children when they realized that their children were not feeling well as shown in the bar graph below.

**Fig 2.**
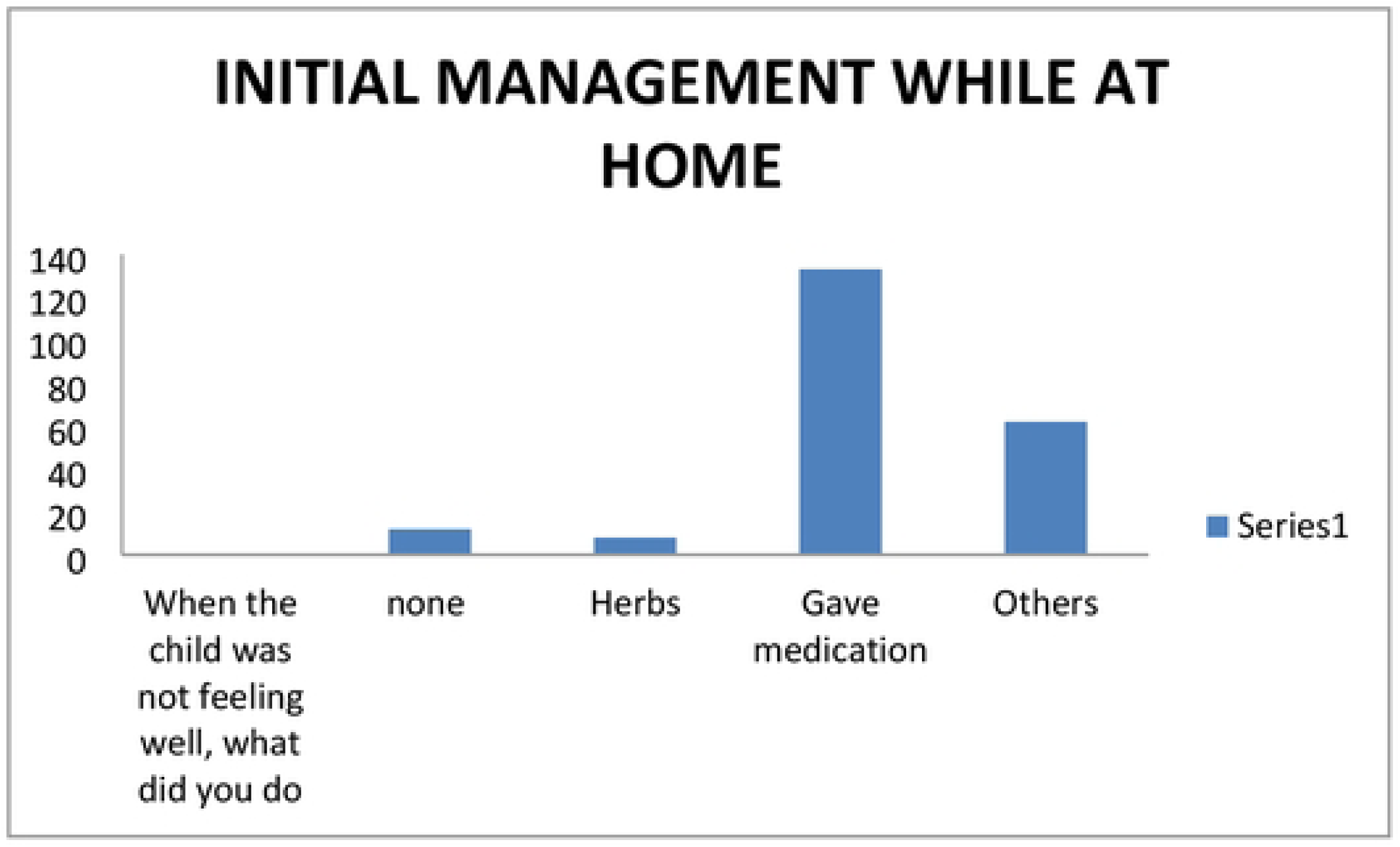
A graph showing the initial management offered to the child while at home.

### Summary of the outcomes

Prior to their admission to the health facility, all of the children presented with fever. Majority of the respondents (83.33%) had spent 1 to 5 days at the facility at the time of data collection. The average number of days spent at the health facility was 3 days. At the point of contact with the child, 92.59% had progressed to severe malaria.

The outcomes of delayed access of care are summarized in the table below,

**Table 3:** The outcomes of delayed access of care.

| VARIABLE |  | FREQUENCY | PERCENTAGE |
| --- | --- | --- | --- |
| Number of days spent at the facility | 1 to 5 days | 180 | 83.33 |
|  | >5 days | 36 | 16.67 |
| manifestations prior to admission | Convulsions | 44 | 20.37 |
|  | Hematuria | 97 | 44.91 |
|  | Fever | 216 | 100 |
|  | Severe anemia | 170 | 78.70 |
|  | Loss of consciousness | 12 | 5.56 |
| Outcomes at time of contact with the child | Progressed to severe malaria | 200 | 92.59 |
|  | Prolonged hospital stay | 37 | 17.13 |
|  | Recovery /responding to treatment | 188 | 87.04 |
|  | Others | 0 | 0 |

### Manifestations of children prior to admission at Mbale Regional Referral Hospital

Most of the children(36%) presented with fever prior to admission. The least number of children (2%) admitted at MRRH presented with loss of consciousness as illustrated on the pie chart below.

**Fig 3.**
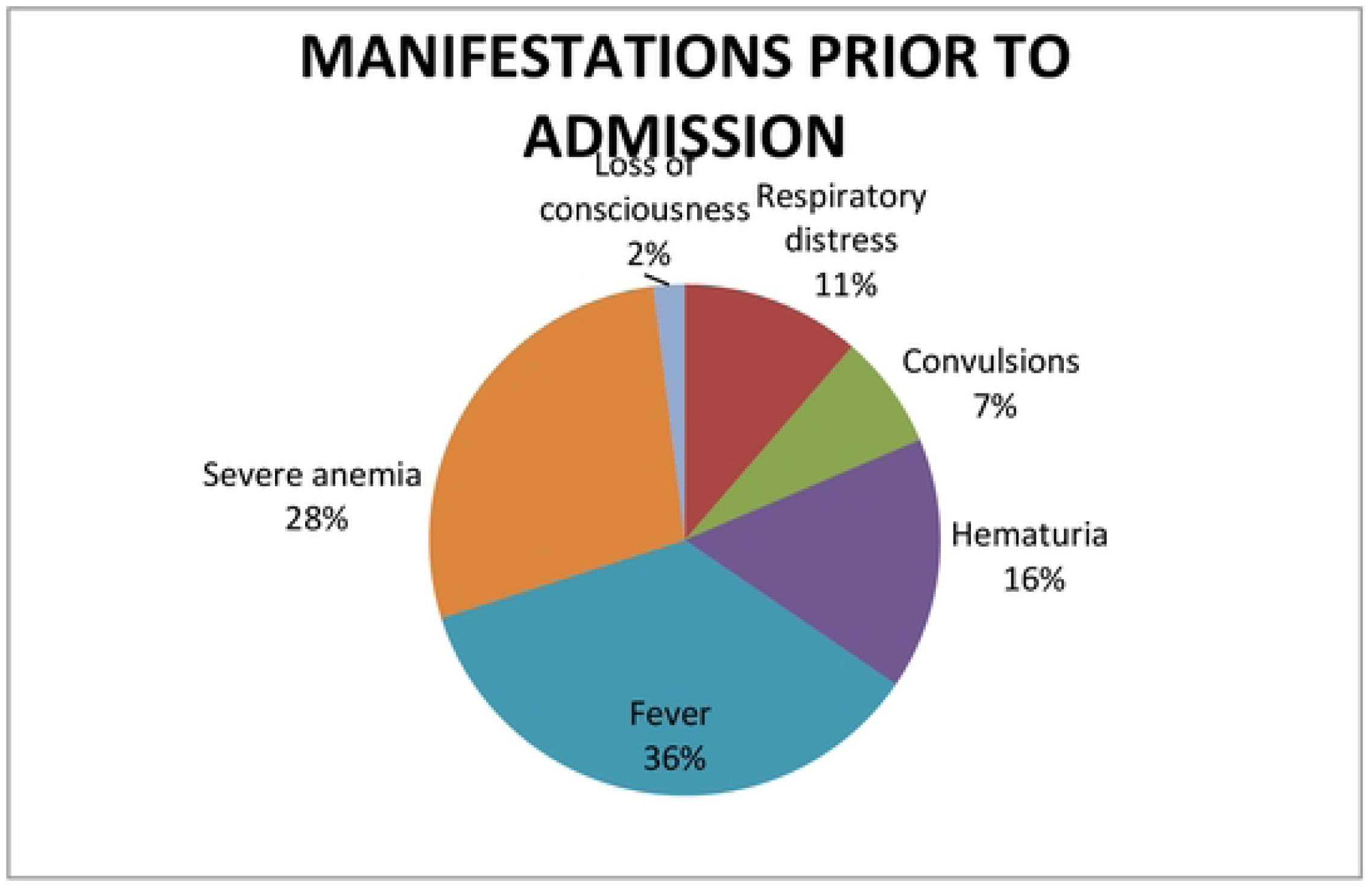
A pie chart illustrating the manifestations that the children had prior to their admission at Mbale Regional Referral Hospital.

### Outcomes of delayed access of care at the point of contact with the child

Most of the children (92.59) had progressed to severe malaria but were reported to be responding to the medication given. However, 17.13% of the children had prolonged hospital stay as shown in the graph below.

**Fig 4.**
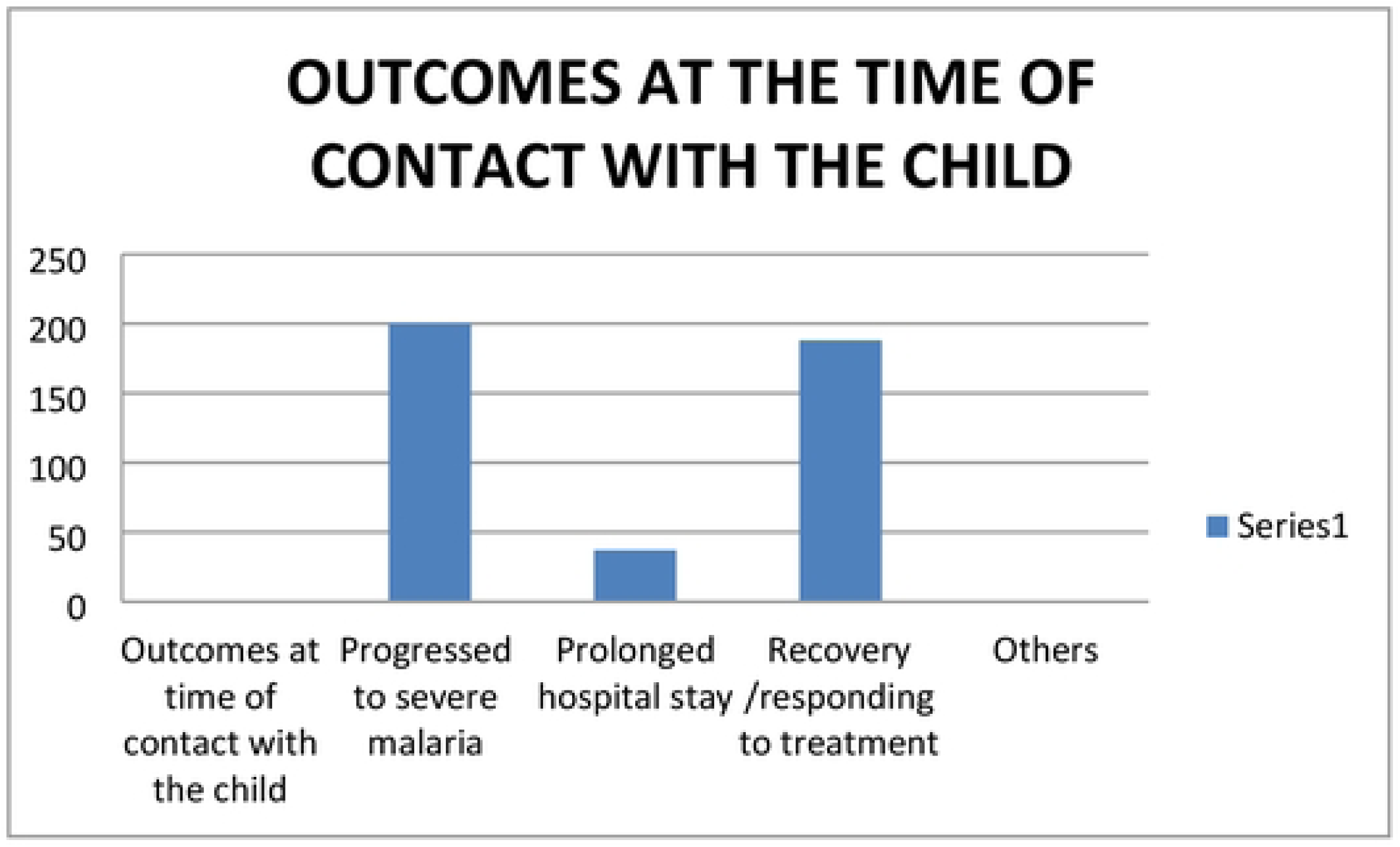
A graph showing the outcomes of delayed access of care at the point of contact with the child.

### Binomial logistic regression between the independent variables and timing of access of care

In bivariate analysis, the caretaker’s highest education level, the initial management of malaria before going to the health facility and the first choice of the health facility were associated with delayed access of care.

The respondents’ level of education was noted most significant. Respondents who had attained tertiary level of education were 5.5 times more likely to delay compared to those with no formal education.

The other significant factors were highlighted in the table below.

**Table 4:**
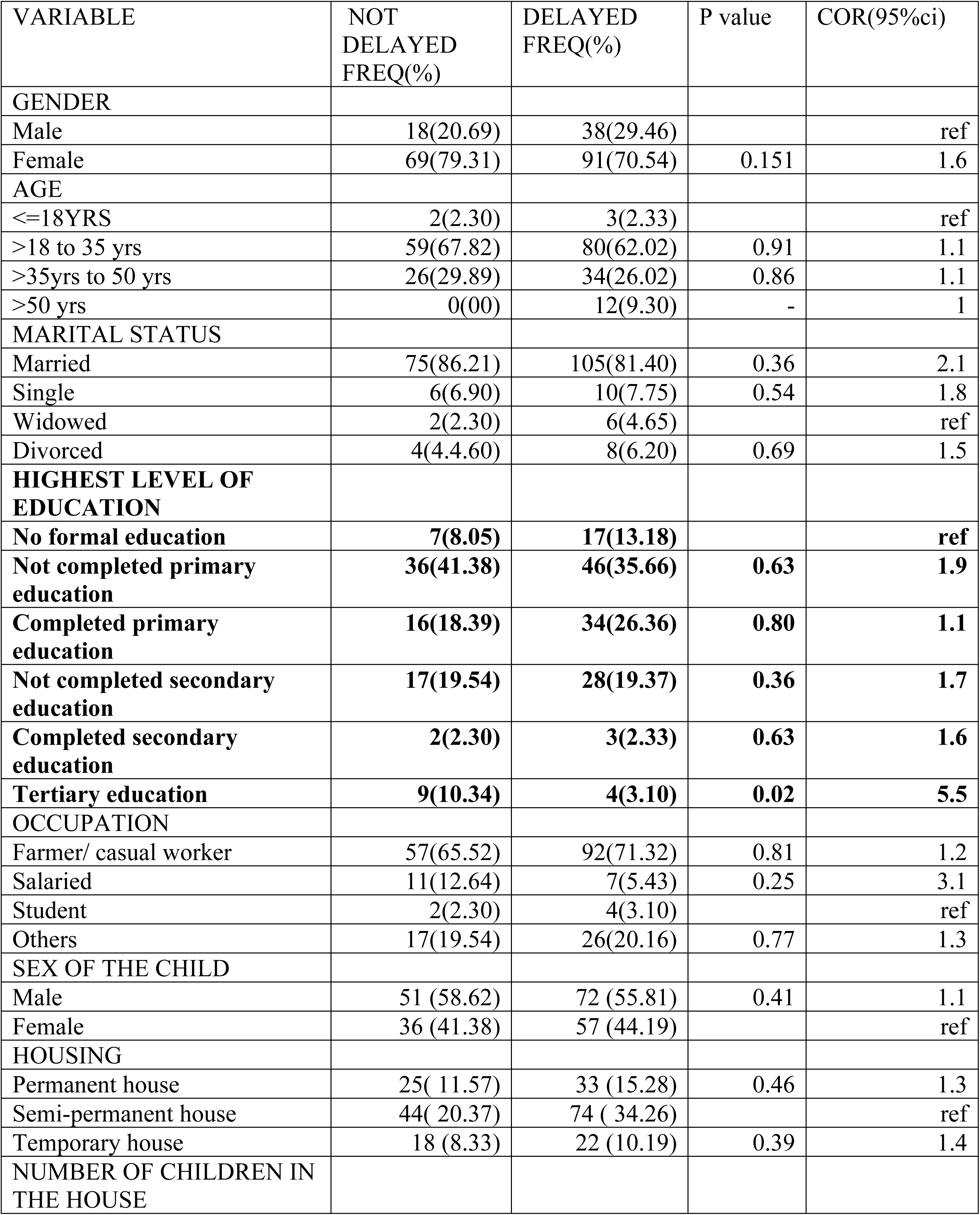
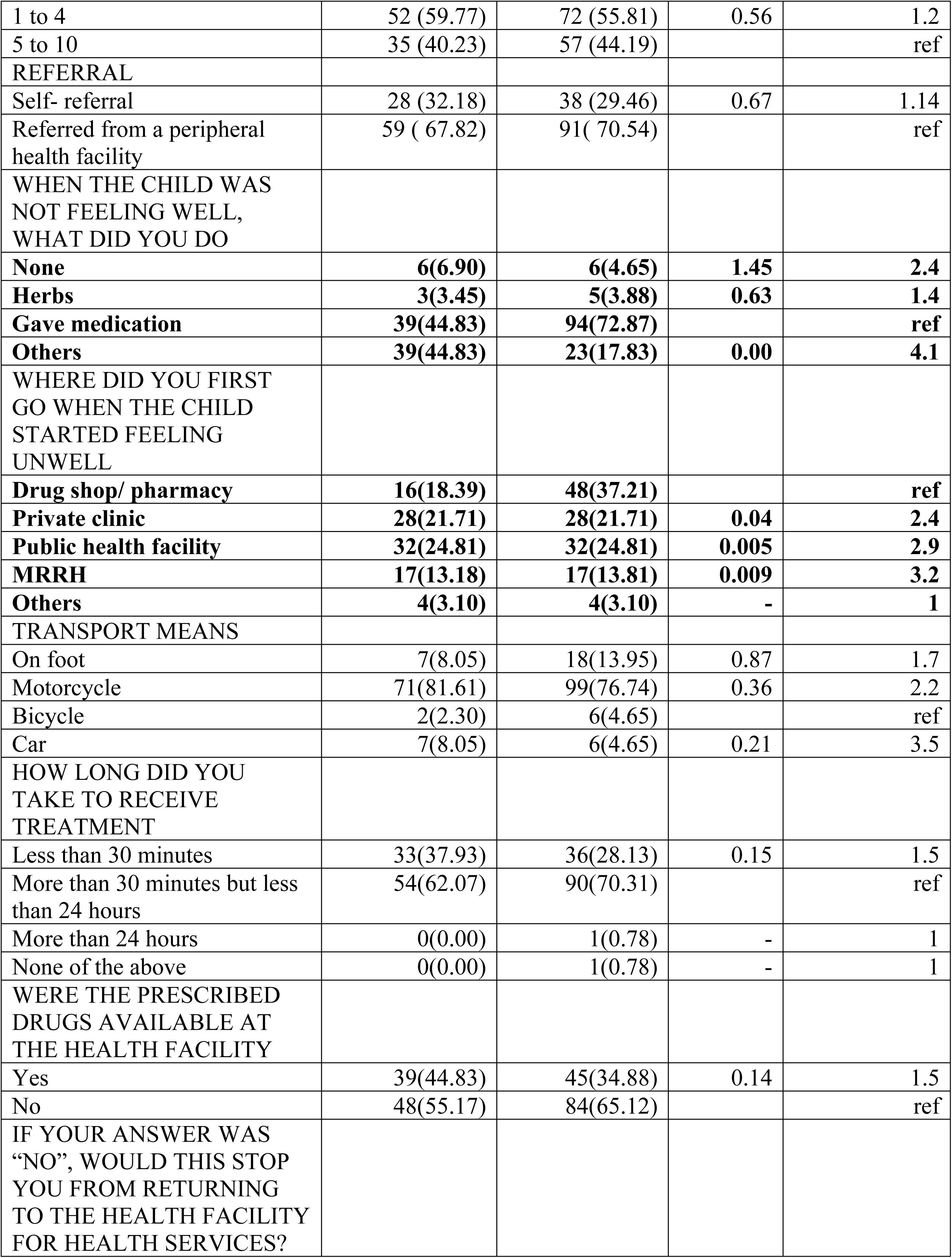
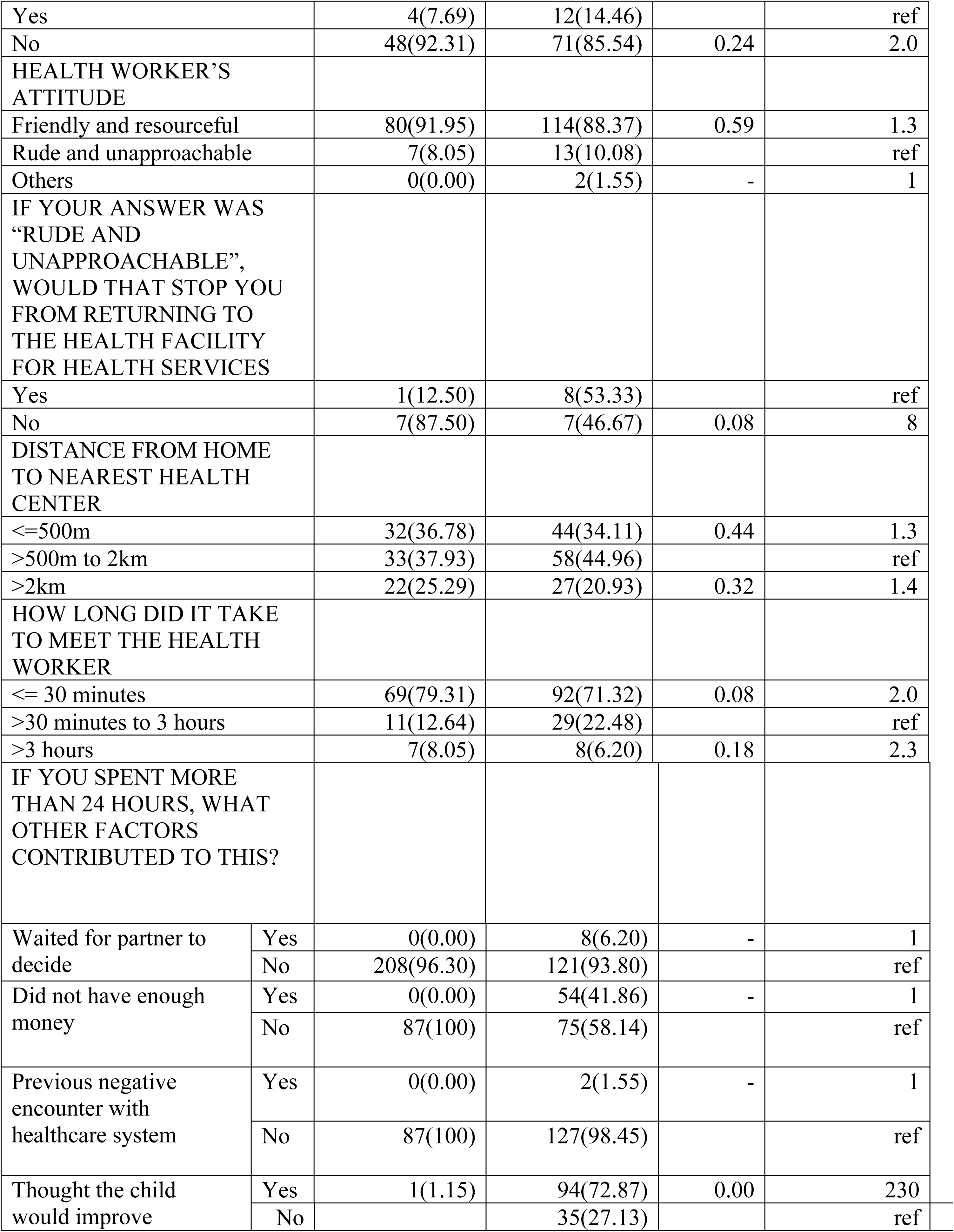
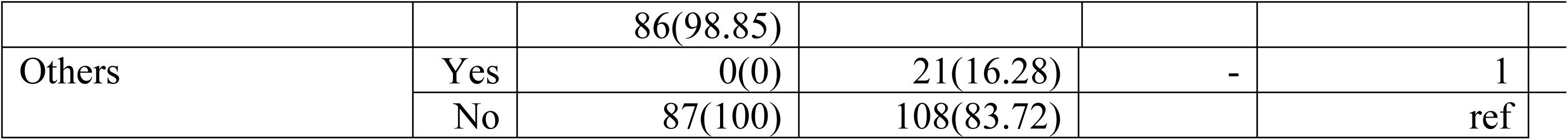
Binomial logistic regression between the independent variables and timing of access of care.

| VARIABLE | NOT<br>DELAYED<br>FREQ(%) | DELAYED<br>FREQ(%) | P value | COR(95%ci) |
| --- | --- | --- | --- | --- |
| **GENDER** |  |  |  |  |
| Male | 18(20.69) | 38(29.46) |  | ref |
| Female | 69(79.31) | 91(70.54) | 0.151 | 1.6 |
| **AGE** |  |  |  |  |
| <=18YRS | 2(2.30) | 3(2.33) |  | ref |
| >18 to 35 yrs | 59(67.82) | 80(62.02) | 0.91 | 1.1 |
| >35yrs to 50 yrs | 26(29.89) | 34(26.02) | 0.86 | 1.1 |
| >50 yrs | 0(00) | 12(9.30) | - | 1 |
| **MARITAL STATUS** |  |  |  |  |
| Married | 75(86.21) | 105(81.40) | 0.36 | 2.1 |
| Single | 6(6.90) | 10(7.75) | 0.54 | 1.8 |
| Widowed | 2(2.30) | 6(4.65) |  | ref |
| Divorced | 4(4.4.60) | 8(6.20) | 0.69 | 1.5 |
| **HIGHEST LEVEL OF**<br>**EDUCATION** |  |  |  |  |
| **No formal education** | **7(8.05)** | **17(13.18)** |  | **ref** |
| **Not completed primary**<br>**education** | **36(41.38)** | **46(35.66)** | **0.63** | **1.9** |
| **Completed primary**<br>**education** | **16(18.39)** | **34(26.36)** | **0.80** | **1.1** |
| **Not completed secondary**<br>**education** | **17(19.54)** | **28(19.37)** | **0.36** | **1.7** |
| **Completed secondary**<br>**education** | **2(2.30)** | **3(2.33)** | **0.63** | **1.6** |
| **Tertiary education** | **9(10.34)** | **4(3.10)** | **0.02** | **5.5** |
| **OCCUPATION** |  |  |  |  |
| Farmer/ casual worker | 57(65.52) | 92(71.32) | 0.81 | 1.2 |
| Salaried | 11(12.64) | 7(5.43) | 0.25 | 3.1 |
| Student | 2(2.30) | 4(3.10) |  | ref |
| Others | 17(19.54) | 26(20.16) | 0.77 | 1.3 |
| **SEX OF THE CHILD** |  |  |  |  |
| Male | 51 (58.62) | 72 (55.81) | 0.41 | 1.1 |
| Female | 36 (41.38) | 57 (44.19) |  | ref |
| **HOUSING** |  |  |  |  |
| Permanent house | 25( 11.57) | 33 (15.28) | 0.46 | 1.3 |
| Semi-permanent house | 44( 20.37) | 74 ( 34.26) |  | ref |
| Temporary house | 18 (8.33) | 22 (10.19) | 0.39 | 1.4 |
| **NUMBER OF CHILDREN IN**<br>**THE HOUSE** |  |  |  |  |
| 1 to 4 | 52 (59.77) | 72 (55.81) | 0.56 | 1.2 |
| 5 to 10 | 35 (40.23) | 57 (44.19) |  | ref |
| REFERRAL |  |  |  |  |
| Self- referral | 28 (32.18) | 38 (29.46) | 0.67 | 1.14 |
| Referred from a peripheral health facility | 59 ( 67.82) | 91( 70.54) |  | ref |
| WHEN THE CHILD WAS NOT FEELING WELL, WHAT DID YOU DO |  |  |  |  |
| **None** | **6(6.90)** | **6(4.65)** | **1.45** | **2.4** |
| **Herbs** | **3(3.45)** | **5(3.88)** | **0.63** | **1.4** |
| **Gave medication** | **39(44.83)** | **94(72.87)** |  | **ref** |
| **Others** | **39(44.83)** | **23(17.83)** | **0.00** | **4.1** |
| WHERE DID YOU FIRST GO WHEN THE CHILD STARTED FEELING UNWELL |  |  |  |  |
| **Drug shop/ pharmacy** | **16(18.39)** | **48(37.21)** |  | **ref** |
| **Private clinic** | **28(21.71)** | **28(21.71)** | **0.04** | **2.4** |
| **Public health facility** | **32(24.81)** | **32(24.81)** | **0.005** | **2.9** |
| **MRRH** | **17(13.18)** | **17(13.81)** | **0.009** | **3.2** |
| **Others** | **4(3.10)** | **4(3.10)** | **-** | **1** |
| TRANSPORT MEANS |  |  |  |  |
| On foot | 7(8.05) | 18(13.95) | 0.87 | 1.7 |
| Motorcycle | 71(81.61) | 99(76.74) | 0.36 | 2.2 |
| Bicycle | 2(2.30) | 6(4.65) |  | ref |
| Car | 7(8.05) | 6(4.65) | 0.21 | 3.5 |
| HOW LONG DID YOU TAKE TO RECEIVE TREATMENT |  |  |  |  |
| Less than 30 minutes | 33(37.93) | 36(28.13) | 0.15 | 1.5 |
| More than 30 minutes but less than 24 hours | 54(62.07) | 90(70.31) |  | ref |
| More than 24 hours | 0(0.00) | 1(0.78) | - | 1 |
| None of the above | 0(0.00) | 1(0.78) | - | 1 |
| WERE THE PRESCRIBED DRUGS AVAILABLE AT THE HEALTH FACILITY |  |  |  |  |
| Yes | 39(44.83) | 45(34.88) | 0.14 | 1.5 |
| No | 48(55.17) | 84(65.12) |  | ref |
| IF YOUR ANSWER WAS “NO”, WOULD THIS STOP YOU FROM RETURNING TO THE HEALTH FACILITY FOR HEALTH SERVICES? |  |  |  |  |

|  |  |  |  |  |  |
| --- | --- | --- | --- | --- | --- |
| Yes |  | 4(7.69) | 12(14.46) |  | ref |
| No |  | 48(92.31) | 71(85.54) | 0.24 | 2.0 |
| HEALTH WORKER'S ATTITUDE |  |  |  |  |  |
| Friendly and resourceful |  | 80(91.95) | 114(88.37) | 0.59 | 1.3 |
| Rude and unapproachable |  | 7(8.05) | 13(10.08) |  | ref |
| Others |  | 0(0.00) | 2(1.55) | - | 1 |
| IF YOUR ANSWER WAS "RUDE AND UNAPPROACHABLE", WOULD THAT STOP YOU FROM RETURNING TO THE HEALTH FACILITY FOR HEALTH SERVICES |  |  |  |  |  |
| Yes |  | 1(12.50) | 8(53.33) |  | ref |
| No |  | 7(87.50) | 7(46.67) | 0.08 | 8 |
| DISTANCE FROM HOME TO NEAREST HEALTH CENTER |  |  |  |  |  |
| <=500m |  | 32(36.78) | 44(34.11) | 0.44 | 1.3 |
| >500m to 2km |  | 33(37.93) | 58(44.96) |  | ref |
| >2km |  | 22(25.29) | 27(20.93) | 0.32 | 1.4 |
| HOW LONG DID IT TAKE TO MEET THE HEALTH WORKER |  |  |  |  |  |
| <= 30 minutes |  | 69(79.31) | 92(71.32) | 0.08 | 2.0 |
| >30 minutes to 3 hours |  | 11(12.64) | 29(22.48) |  | ref |
| >3 hours |  | 7(8.05) | 8(6.20) | 0.18 | 2.3 |
| IF YOU SPENT MORE THAN 24 HOURS, WHAT OTHER FACTORS CONTRIBUTED TO THIS? |  |  |  |  |  |
| Waited for partner to decide | Yes | 0(0.00) | 8(6.20) | - | 1 |
|  | No | 208(96.30) | 121(93.80) |  | ref |
| Did not have enough money | Yes | 0(0.00) | 54(41.86) | - | 1 |
|  | No | 87(100) | 75(58.14) |  | ref |
| Previous negative encounter with healthcare system | Yes | 0(0.00) | 2(1.55) | - | 1 |
|  | No | 87(100) | 127(98.45) |  | ref |
| Thought the child would improve | Yes | 1(1.15) | 94(72.87) | 0.00 | 230 |
|  | No |  | 35(27.13) |  | ref |
|  |  | 86(98.85) |  |  |  |
| Others | Yes | 0(0) | 21(16.28) | - | 1 |
|  | No | 87(100) | 108(83.72) |  | ref |

### Binomial logistic regression between the outcomes of delay and timing of access of care

None of the outcomes of delayed access of care was associated with delayed access of care.

The binomial logistic regression between the outcomes of delayed access of care (number of days spent at the health facility, manifestations prior to admission and outcomes at the point of contact with the child) and delayed access of care are summarized below.

**Table 5:** Binomial logistic regression between the outcomes of delay and timing of access of care.

| VARIABLE |  | NOT DELAYED<br>FREQ (%) | DELAYED<br>FREQ (%) | P value | COR (95%<br>ci) |
| --- | --- | --- | --- | --- | --- |
| NUMBER OF DAYS<br>SPENT AT THE<br>FACILITY |  |  |  |  |  |
| 1 to 5 days |  | 75(86.21) | 104(80.62) | 0.29 | 1.5 |
| >5 days |  | 12(13.79) | 25(19.38) |  | ref |
| MANIFESTATIONS<br>PRIOR TO<br>ADMISSION |  |  |  |  |  |
| Respiratory<br>distress | Yes | 37(42.53) | 61(47.29) |  | ref |
|  | No | 50(57.47) | 68(52.71) | 0.49 | 1.2 |
| Convulsions | Yes | 28(32.18) | 32(24.81) | 0.24 | 1.4 |
|  | No | 59(67.82) | 97(75.19) |  | ref |
| Hematuria | Yes | 48(55.17) | 77(59.69) | 0.52 | 0.8 |
|  | No | 39(44.83) | 52(40.31) |  | ref |
| Fever | Yes | 87(100) | 129(100) | - | 1 |
|  | No | - | - | - | ref |
| Severe anemia | Yes | 83(95.40) | 119(92.25) |  | ref |
|  | No | 4(4.60) | 10(7.75) | 0.16 | 1.6 |
| Loss of<br>consciousness | Yes | 10(11.49) | 13(89.92) | 0.74 | 1.2 |
|  | No | 77(88.51) | 116(89.92) |  | ref |
| Outcomes at time of contact with the child |  |  |  |  |  |
| Progressed to severe malaria | Yes | 83(95.40) | 119(92.25) | 0.36 | 1.7 |
|  | No | 4(4.60) | 10(7.75) |  | ref |
| Prolonged hospital stay | Yes | 11(12.64) | 30(23.26) |  | ref |
|  | No | 76(87.36) | 99(76.74) | 0.054 | 2.1 |
| Recovery /responding to treatment | Yes | 77(88.50) | 114(88.37) | 0.98 | 1.0 |
|  | No | 10(11.49) | 15(11.63) |  | ref |

### 4.6 Binomial logistic regression between the delayed access of care and outcomes of timing of access of care

When delay is run as the independent variable against the outcomes, time taken to seek care is not associated with outcomes like severe malaria, prolonged hospital stay and recovery as summarized in the table below.

**Table 6:** Binomial logistic regression between the delayed access of care and outcomes of timing of access of care.

|  | OUTCOMES |  |  |  |  |  |  |  |  |  |  |  |
| --- | --- | --- | --- | --- | --- | --- | --- | --- | --- | --- | --- | --- |
|  | SEVERE MALARIA |  | P value | C O R | PROLONGED HOSPITAL STAY |  | P value | C O R | RECOVERY |  | P value | C O R |
|  | YES | NO |  |  | YES | NO |  |  | YES | N O |  |  |
| DELAYED | 119(58.91) | 10(71.43) |  | ref | 30(73.17) | 99(56.57) | 0.054 | 2.09 | 114(59.72) |  |  | ref |
| DID NOT DELAY | 83(41.09) | 4(28.57) | 0.36 | 1.7 | 11(26.83) | 76(43.43) |  | ref | 77(40.31) |  | 0.98 | 1.01 |

#### Multinomial logistic regression between independent variables and timing of access of care

In multivariate analysis, the highest education level had a very significant association with delayed access of care and tertiary education had a more significant association with a p value of 0.02. Initial management when the child had fallen ill had a significant association with delayed access of care. Other options of initial management when the child was ill had a significant association with delayed access of care with a p value of 0.00.

**Table 7:** The multinomial logistic regression between independent variables and timing of access of care.

| Variable | Category | P value | AOR |
| --- | --- | --- | --- |
| Education | Did not complete primary education | 1.20 | 2.0 |
|  | Completed primary education | 0.90 | 1.1 |
|  | Did not complete secondary education | 0.44 | 1.6 |
|  | Completed secondary education | 0.83 | 1.3 |
|  | Tertiary education | 0.02 | 7.1 |
|  | No formal education |  | ref |
| When the child was not feeling well, what did you do | Herbs | 0.60 | 1.5 |
|  | Gave medication |  | ref |
|  | Others | 0.00 |  |
|  | None | 0.21 | 2.2 |
| Where did you first go when the child started feeling unwell | MRRH | 0.23 | 1.8 |
|  | Private facility | 0.46 | 1.4 |
|  | Public health facility | 0.23 | 1.7 |
|  | Drug shop/ pharmacy |  | ref |
|  | others | - | 1 |

## Discussion

This study found that the highest level of education of the caretaker and being treated at home before getting to the health facility were associated with delayed access of care. This is similar to a study in Bata(3).

### Socio-demographic characteristics

In this study, 74% of the respondents were female most likely due to the fact that nurturing of children is regarded to be a female’s role.

Of the caretakers who delayed to seek care 62.02% were between the ages of 18 and 35. A study in Ethiopia associated younger maternal age to timely seeking of care when a child fell ill(19) which wasn’t the case in this study. There was no association between parental/caretaker’s age and delayed access of care probably due to the fact that many other factors are considered before one decided to seek care for an ill child. The decision to seek care when a child fell ill was usually left to the head of the family(15) while others consulted elderly family members who seemed more knowledgeable on the next course of action(10).

Caretakers who had attained tertiary education were 7.1 times more likely to delay compared to those with no formal education(AOR=7.2, p value=0.02) which was probably due to the fact that they spent most of their time at their work places away from home. Another study in Malawi however found that there was no association between prompt seeking of care and the care giver’s level of education(20). A study in Kaduna showed that delay in seeking care for fever among children U5 was strongly associated with no formal education of the caregiver(21).

Delays in seeking care are linked to a low economic status(22) which wasn’t the case in this study despite the fact that 61.7% of the 149 who were casual laborers delayed to seek care. This is most likely due to the fact that the cost of healthcare varies depending on the choice of the health facility and the availability of free drugs at that facility.

The more the number the number of children in a household, the higher the likelihood of delayed access of care when they fell ill(23). This wasn’t the case in this study where there was no association between the number of children in the household and delayed access of care. This is probably due to the fact that the study participants were under the age of 5 and caretakers tend to prioritize their health and wellbeing

Timely seeking of care when ill was associated with the child being male(24) which wasn’t the case in this study where there was no association between the sex of the child and delayed access of care. This is most probably due to the reduction in gender disparities over time.

### Delays

Majority of the respondents (59.26%) reported to have taken more than 24 hours to reach the health facility when their children fell ill and attributed it to the lack of money (24.54%), the thought that their children would improve (43.52%), previous negative encounter with the healthcare system (0.46%) and other factors like their absence at the time when the child was ill (10.19%). Only 3.70% of those who delayed to access care attributed it to the fact they had been waiting for their partner to make that decision. The average time taken to seek care was 1 day and 6 hours with a median of 1 day and a mode of 2 days. Majority of those who had thought that their children would improve had initially administered medication to their ill children before visiting a hospital.

The initial use of old prescriptions, leftover drugs(25)(25) and herbal medicine(26) further led to delayed access of care. In this study, 61.57% of the respondents reported to having given their children medication obtained from drug shops and others were left over drugs. The drugs given included ACTs and paracetamol which most respondents had considered to be first aid treatment. These drugs were given to manage fever, which all the children in this study were reported to have had and other signs that the respondents believed to be consistent with Malaria since it was a routine illness. Those who initially implemented measures other than giving medication/herbs before taking a child to the health center were 4.1 time more likely to delay compared to those who had initially given their children medication before reaching the health facility(AOR=4.1, p value=0.00). The other measures included monitoring the child to see if they would improve and taking time while sourcing for resources to take the child to the health center among others. More than half of the respondents (52.78%) initially opted for private facilities before coming to MRRH. These findings are similar to those of a study done in Uganda whereby drug shops were preferred to government facilities that were considered to be expensive and distant(10).

Distance to the health facility is not always associated with the delay in access of care(20). These findings are consistent with those found in this study.

Drug stock out(11), long waiting hours and a previous negative encounter with health workers(27) were associated with delayed access of care. This study however, found out that long waiting hours, drug stock out and the attitude of the health worker were not always associated with delayed access of care probably due to the fact that the caretakers prioritized their children’s wellbeing and were willing to overlook the prevailing challenges.

### Outcomes of delayed access of care

This study did not find a significant association between manifestations consistent with severe malaria and delayed access of care. This is most likely due to the fact that disease progression varies among individuals. Malaria was only considered to be life threatening when it is associated with anemia, convulsions(28), persistent fever and convulsions(29) which then prompted timely management.

Adverse health outcomes like mortality, prolonged hospital stay and development of complications are directly associated with delay in seeking health services when ill(30). Timely therapeutic interventions are required to prevent such outcomes(31). A meta-analysis on the impact of delayed treatment of uncomplicated *P falciparum* malaria on progression to severe malaria suggested that about 50% of the severe malaria cases would have been prevented if antimalarial treatment had been started within 24hrs of symptom onset(32).

Findings from a meta-analysis carried out in selected African countries including Uganda showed that mortality following admission depended on the severity of the disease where 1% of the study participants died due to uncomplicated malaria and 8.3% due to severe malaria. The deaths were further associated with delay in receiving blood transfusions among those with severe malaria and poor long term outcomes among those who survived(32). Most of the respondents (69.44%) reported to having been referred from peripheral facilities to MRRH due to respiratory distress and a low hemoglobin level that required blood transfusion among others.

An increase in in-patient length of stay was associated with admission delay at a hospital in Canada(33). This study however, showed that the association between outcomes of delayed access of care (progression to severe malaria, prolonged hospital stay and recovery) and the delay to access care is not always significant. This is most likely due to the fact that delayed access of care is multifactorial and the concept of delay varies among individuals basing on the signs they manifest with.

### Limitations

The sample used in this study is biased towards the most ill children hence affecting the generalizability of the findings.

The study was conducted among parents/caretakers of children attending one health facility which may limit the generalizability of the findings to all caretakers in the community. In addition, because the study design was cross-sectional, causal inferences could not be made.

## Conclusion

Despite the numerous interventions put into place to curb the spread of malaria and to manage malaria, the issue of delayed access of care remains a significant contributor to the adverse effects of malaria among children under five.

## Recommendations

Health education on the impact of delayed access of care should be intensified at all levels of healthcare. This is because health seeking among children under five is entirely dependent upon their parents/caretakers.

## Data Availability

Data from this study will be made available by the corresponding authors on a reasonable request.

## Acknowledgements

We acknowledge the support from staff at Mbale regional referral pediatric and acute wards during the data collection process for this study.

## Author Contributions

**Conceptualization:** Kayeny Miriam Melody Yung, Pamella R Adongo

**Data curation:** Kayeny Miriam Melody Yung

**Formal Analysis:** Kayeny Miriam Melody Yung

**Methodology:** Kayeny Miriam Melody Yung

**Supervision:** Pamella R Adongo

**Validation and visualization**: Kayeny Miriam Melody Yung

**Validation:** Pamella R Adongo

**Writing – original draft:** Kayeny Miriam Melody Yung

**Writing – review & editing:** Ivan Lyagoba, Lydia VN Ssenyonga, Paul Oboth, Samuel Olowo

## Notes

### Competing Interest Statement

The authors have declared no competing interest.

### Author Declarations

Ethical procedures were all met, with approval obtained from Mbale Regional Referral Research and Ethics Committee (UG-REC-011) under the reference number MRRH-2022-208.

